# Preclinical and first-in-human-brain-cancer applications of [18F]poly-(ADP-ribose) polymerase inhibitor PET/MR

**DOI:** 10.1101/2020.07.13.20141036

**Authors:** Robert J Young, Paula Demétrio De Souza França, Giacomo Pirovano, Anna F Piotrowski, Philip J Nicklin, Christopher C Riedl, Jazmin Schwartz, Tejus A Bale, Patrick L Donabedian, Susanne Kossatz, Eva M Burnazi, Sheryl Roberts, Serge K Lyashchenko, Alexandra M Miller, Nelson S. Moss, Megan Fiasconaro, Zhigang Zhang, Audrey Mauguen, Thomas Reiner, Mark P Dunphy

**Affiliations:** Department of Radiology, Memorial Sloan Kettering Cancer Center, New York, New York; Department of Otorhinolaryngology and Head and Neck Surgery, Federal University of São Paulo, SP, Brazil; Department of Neurology, Memorial Sloan Kettering Cancer Center, New York, New York; Department of Medical Physics, Memorial Sloan Kettering Cancer Center, New York, New York; Department of Pathology, Memorial Sloan Kettering Cancer Center, New York, New York; Department of Neurosurgery and Brain Metastasis Center, Memorial Sloan Kettering Cancer Center, New York, New York; Department of Biostatistics and Epidemiology, Memorial Sloan Kettering Cancer Center, New York, New York; The Brain Tumor Center, Memorial Sloan Kettering Cancer Center, New York, New York; Weill Cornell Medical College, New York, NY, USA; Chemical Biology Program, Memorial Sloan Kettering Cancer Center, New York, NY, USA; Department of Radiology, Weill Cornell Medical College, New York, NY, USA

## Abstract

We report pre-clinical and first-in-human-brain-cancer data using a targeted poly(ADP-ribose)polymerase1 (PARP1) binding PET tracer, [^18^F]PARPi, as a diagnostic tool to differentiate between brain cancers and treatment related changes. In a pre-clinical mouse model, we illustrated that [^18^F]PARPi crosses the blood-brain barrier and specifically binds to PARP1 overexpressed in cancer cell nuclei. In humans, we demonstrated high [^18^F]PARPi uptake on PET/MR in active brain cancers and low uptake in treatment related changes, independent of blood brain-barrier disruption. Immunohistochemistry results confirmed higher PARP1 expression in cancers than non-cancers. Specificity was also corroborated by blocking fluorescent tracer uptake with excess of unlabeled PARP inhibitor in fresh cancer tissue derived from a patient. Although larger studies are necessary to confirm and further explore this tracer, we describe an encouraging role for the use of [^18^F]PARPi as a diagnostic tool in evaluating patients with brain cancers and possible treatment related changes.

**One Sentence summary:** PET imaging with [^18^F]PARPi can differentiate active brain cancer from treatment related changes with encouraging results for use during treatment follow-up.

## INTRODUCTION

Positron emission tomography (PET) scans of the brain are often performed to identify and distinguish cancers from infection and from other non-cancer entities at diagnosis, most importantly from alterations related to prior treatment. Currently, [^18^F]fluorodeoxyglucose (FDG) is the only radiotracer approved in the United States by the Food and Drug Administration (FDA); however, its sensitivity and specificity are known to be limited due to the high glucose uptake of the normal brain and also prominent uptake with postoperative and treatment related inflammatory changes (*1, 2*). Because of the high physiologic uptake in normal gray matter, FDG-avid brain cancers are also often indistinct on PET brain scans.

In Europe, amino acid PET radiotracers, in particular [^18^F]fluoroethyltyrosine ([^18^F]FET), are widely available and preferred for brain cancer imaging as normal brain tissues demonstrate less uptake of the amino acid tracer when compared to the glucose tracer, improving the cancer-to-background contrast (*3*). A recent meta-analysis indicates that [^18^F]FET PET is probably superior to [^18^F]FDG PET in differentiating between brain cancer progression versus treatment related changes, although the 95% confidence intervals (CI) overlapped: [^18^F]FET PET had pooled sensitivity of 90% (95% CI, 81-95%) and specificity of 85% (95% CI, 71-93%) versus [^18^F]FDG PET with pooled sensitivity of 84% (95% CI, 72-92%) and specificity of 84% (95% CI, 69-93%) (*4*). Nevertheless, the limited clinical accuracy of current brain PET tracers clearly indicates the need for new diagnostic agents.

We investigate the use of fluorine-18 labeled poly (ADP-ribose) polymerase1 inhibitor ([^18^F]PARPi) as a potentially useful technique to image brain cancers. In a preclinical study with the head-to-head comparison of [^18^F]FET and [^18^F]PARPi in a mouse U251 xenograft model, [^18^F]PARPi demonstrated superior cancer visualization and superior lesion-to-contralateral uptake ratios (*5*). Unlike FDG, cancer detection with [^18^F]PARPi is not based on metabolic activity but rather on the presence of the DNA-repair enzyme poly (ADP-ribose) polymerase1 (PARP1) inside the cancer cell nuclei (*6, 7*). The poly (ADP-ribose) polymerase (PARP) family of DNA repair enzymes is overexpressed in many solid cancers including brain metastases and high grade gliomas (*8-10*). This overexpression is thought to represent a cellular response to the genomic instability and the frequent cell division occurring in cancer cells (*11*). The difference in PARP1 expression found in between cancer and normal tissue generates the contrast that can be seen on a PET scan after injection of [^18^F]PARPi.

The [^18^F]PARPi radiotracer is structurally similar to the PARP inhibitor olaparib (AstraZeneca, Cambridge, UK), and its ability to target the PARP1 enzyme within the cell nucleus is maintained since the structural modification of the drug is made on the cyclopropane end of the olaparib scaffold, a change that doesn’t perturb target binding (*12*). In pre-clinical work, [^18^F]PARPi has been shown to visualize glioblastoma xenograft and orthotopic glioblastoma with 45x greater uptake than in the healthy brain of mouse models (*13*). It has also shown 2x greater uptake in intracranial U251 xenograft cancers as compared to experimentally induced radiation necrosis (*5*).

This pilot study was performed to examine the feasibility of [18F]PARPi imaging in patients with brain cancers and treatment related changes. We hypothesized that active brain cancers will have high [^18^F]PARPi uptake due to overexpression of PARP1/2 in the cancer cells.

## RESULTS

### Mouse

#### [^18^F]PARPi and (FITC)-Dextran have non-overlapping uptake in a glioma mouse model

After intracranial diffuse intrinsic pontine glioma (DIPG) cancers were grown in mice, the animals were co-injected intravenously with 150–170 µCi [^18^F]PARPi and fluorescein isothiocyanate (FITC)-Dextran (Fig. 1 A and B). Mice brains were harvested 1-hour post injection, sliced and imaged. Adjacent slides showed undetectable FITC fluorescence where autoradiography of the adjacent slides presented [^18^F]PARPi signal (Fig. 1 C, top and bottom, respectively), suggesting penetration of [^18^F]PARPi into areas which are inaccessible for the blood-brain barrier impermeable (FITC)-Dextran.

**Fig 1.**
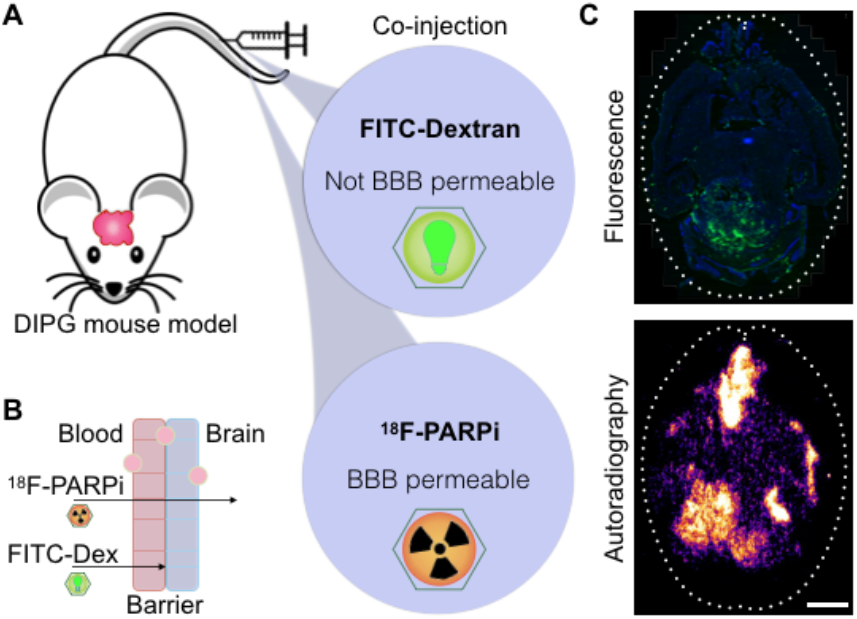
Mouse models demonstrate blood brain barrier permeability of [^18^F]PARPi. (A) A DIPG cancer model was grown in mice for 4-5 weeks. Animals were co-injected (via the tail vein) with 150 – 170 µCi of [^18^F]PARPi and FITC-Dextran (10 kDa). (B) By analyzing the localization of FITC-Dextran, a substance that does not penetrate the blood-brain barrier in normal conditions, and [^18^F]PARPi post injection we demonstrated that an intact blood-brain barrier is able to block dextran passage while allowing some [^18^F]PARPi to pass. (C) Mouse brains were harvested 1 hour post injection, sliced and imaged. Adjacent slides showed undetectable FITC fluorescence where autoradiography of the same slides presented [^18^F]PARPi signal. Scale bar corresponds to 250 µm.

### Human

#### Patient distribution

A total of five patients with seven enhancing lesions ≥1.5 cm were prospectively enrolled onto the study over a four-month period (December 2019 - March 2020), as summarized in Figure 2. The median age of the patients was 49 years old (range, 34-56) and 80% were male. Patient data are summarized in Table 1 (briefly, three patients had isocitrate dehydrogenase (IDH)-wildtype primary glioblastomas and two patients had brain metastases - one with melanoma and three lesions, and one with renal cell carcinoma). Four of the seven lesions (57%) were completely resected after median 2.5 days (range, 1-31) after [^18^F]PARPi positron emission tomography – magnetic resonance (PET/MR). Three lesions were histologically confirmed as cancer: one new untreated glioblastoma and two recurrent metastases. Four lesions were treatment related changes: one was completely resected and showed no cancer; and three were diagnosed based on standard of care clinical and imaging follow up.

**Table 1.**
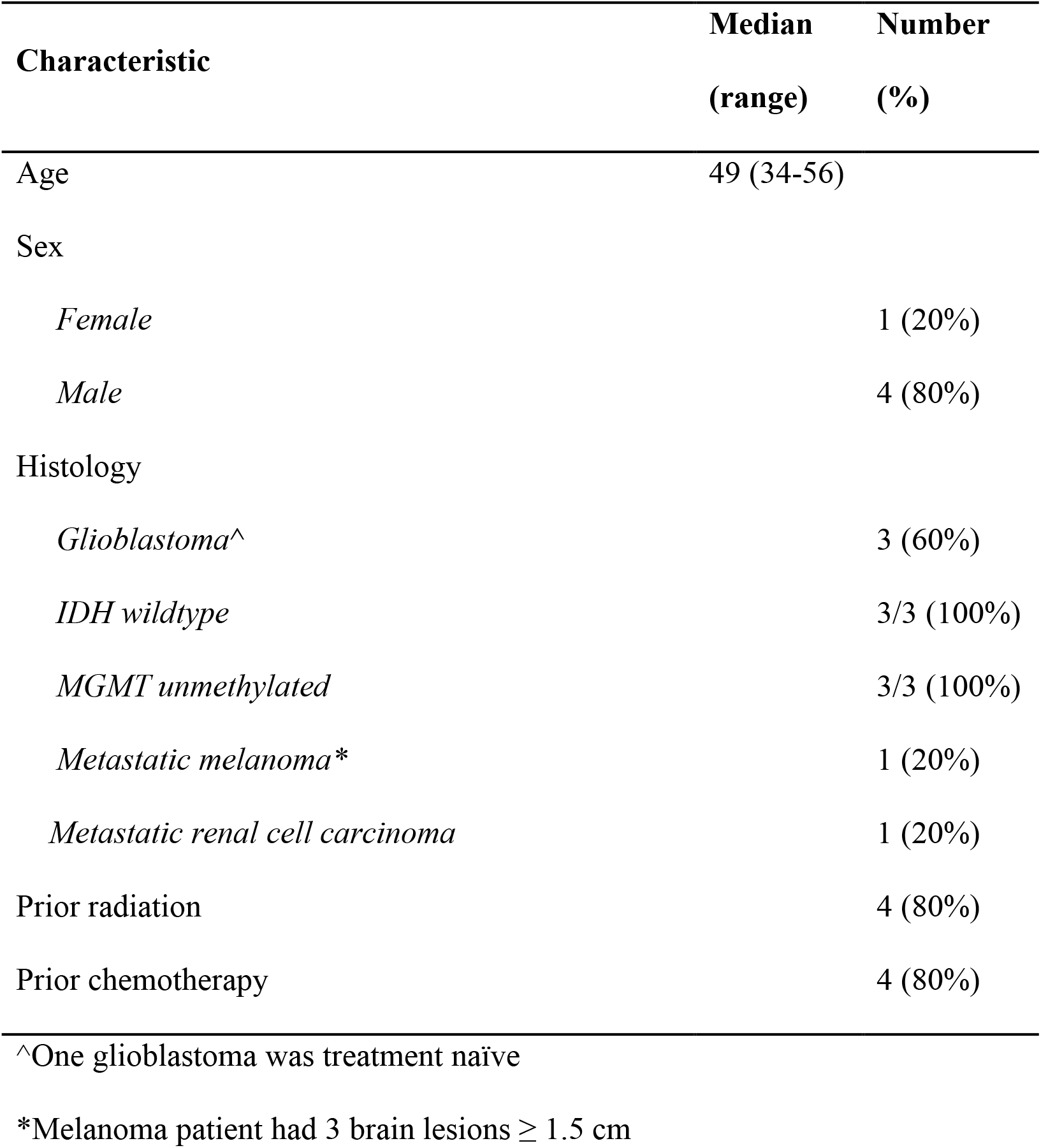
Summary of patient characteristics. Additional alterations in the glioblastomas determined using next-generation sequencing: The glioblastoma showing treatment related changes (patient #1) had PTEN, TERT and FLT4 mutations. The recurrent glioblastoma (patient #3) had TP53, NF1 and PIK3R1 mutations and RB1 deletion. The treatment naïve glioblastoma (patient #5) had TP53, MET and TERT mutations, CDK4 and GLI1 amplifications, and PARK2 and TET2 loss.

**Fig 2.**
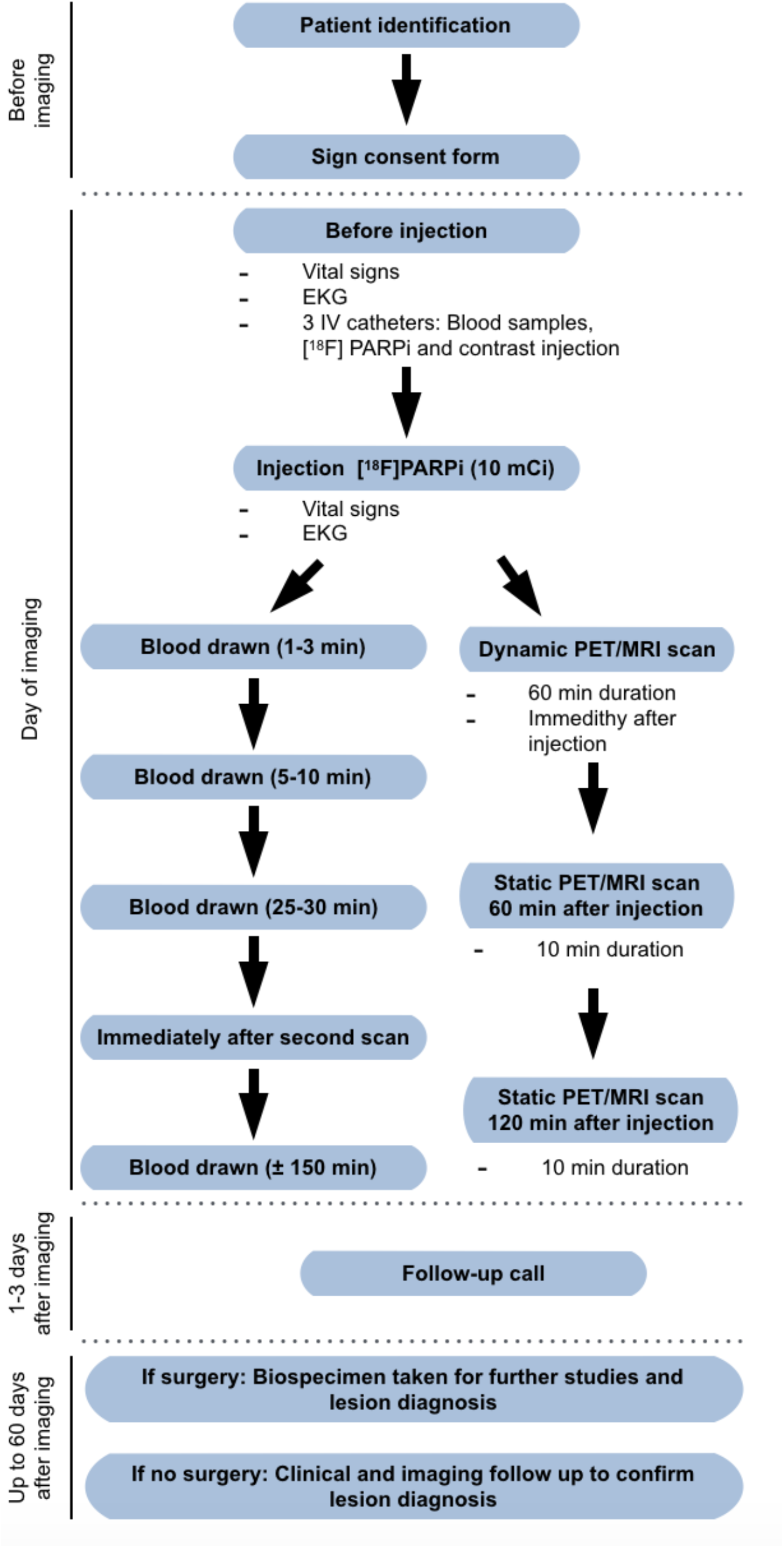
Representation of the study schema. Potential subjects were identified by a member of the patient’s treatment team and referred to the investigator and staff of the study. Participation in the study was voluntary. Intervention consisted of one PET/MR examination with up to 3 scan times using [^18^F]PARPi as a tracer (10 mCi). Six blood draws (5 mL each) were taken: 5 to access the pharmacokinetics and distribution of the drug and one for complete blood count with differential and a complete metabolic panel to access toxicity. Vital signs included temperature, heart rate, blood pressure, oxygen saturation; none were out of normal range.

Patient #1 with glioblastoma was 6 months status post radiation therapy at the time of [^18^F]PARPi PET/MR. Previous treatments included temozolomide, PARP1/2 inhibitor, combination EGFR variant III and CD3 immunotherapy; subsequent treatments included bevacizumab, carboplatin and pembrolizumab. Follow up scan, around 4 months, was consistent with treatment related changes (stable disease by modified RANO). Patient #2 had 3 hemorrhagic enhancing lesions that were around 20, 15 and 10 months status post stereotactic radiosurgery (2100 cGy each). One lesion was resected and diagnosed as recurrent metastasis, and two lesions after follow up were diagnosed as treatment related changes (both stable disease by RANO-BM), although she died from systemic progression 2.5 months after [^18^F]PARPi PET/MR.

#### High [^18^F]PARPi uptake on PET/MR correlated with active cancer lesions

[^18^F]PARPi imaging findings in cancer and treatment related changes are summarized in Table 2. Despite small cohort sizes, the median measurements of standardized uptake value (SUV) at 60 minutes (SUV_60mean_) and the ratio of lesion-to-normal (ratio SUV_60mean_) were higher in the cancer group (1.16 and 1.98, respectively) than in the treatment change group (0.45 and 0.72, respectively, p = 0.03). There was high correlation between the measurements at 60 minutes and 120 minutes, with the latter also increased in cancers with p = 0.03. In all lesions, the K_trans_ and plasma volume (VP) perfusion measurements were also higher in cancers than in treatment related changes (p = 0.03). The contrast clearance analysis trended toward higher values in cancers with p = 0.08. The volume of the enhancing lesion was not different between groups with p = 0.70. Results for individual lesions are reported in Supplementary Table 1.

**Table 2.** Summary of [^18^F]PARPi imaging data.

| Imaging | Cancer | Treatment related change | P-value |
| --- | --- | --- | --- |
|  | n=3<sup>1</sup> | n=4<sup>1</sup> | *Significant |
| [$^{18}\text{F}$]PARPi SUV<sub>60mean</sub> | 1.16 (1.14, 1.19) | 0.45 (0.39, 0.55) | 0.03* |
| [$^{18}\text{F}$]PARPi SUV<sub>120mean</sub> | 0.80 (0.76, 0.82) | 0.34 (0.29, 0.45) | 0.03* |
| [$^{18}\text{F}$]PARPi ratio SUV<sub>60mean</sub> | 1.98 (1.88, 2.00) | 0.72 (0.65, 0.76) | 0.03* |
| [$^{18}\text{F}$]PARPi ratio SUV<sub>120mean</sub> | 1.86 (1.69, 2.10) | 0.95 (0.80, 1.05) | 0.03* |
| Other advanced imaging |  |  |  |
| *Enhancing volume (cm<sup>3</sup>)* | 58 (48, 66) | 80 (42, 117) | 0.7 |
| *DCE, rK<sub>trans</sub>* | 7.20 (6.10, 8.57) | 2.21 (1.73, 2.58) | 0.03* |
| *DCE, rVP* | 4.28 (3.82, 6.16) | 1.49 (1.41, 1.65) | 0.03* |
| *Cancer ratio, CCA* | 0.67 (0.58, 0.73) | 0.38 (0.36, 0.43) | 0.08 |

<sup>1</sup>Statistics presented: median (interquartile range, IQR)
SUV<sub>60mean</sub> = standardized uptake value 60 minutes after injection
SUV<sub>120mean</sub> = standardized uptake value 120 minutes after injection
DCE = dynamic contrast enhanced T1 perfusion
rK<sub>trans</sub> = ratio transfer coefficient constant lesion/normal (measure of leakiness)
VP = ratio plasma volume lesion/normal (measure of perfusion)
CCA = contrast clearance analysis

#### Lesions with high [^18^F]PARPi uptake on scans also demonstrated high PARP1 expression in tissue specimens

PARP1 expression of all resected lesions are represented in Figure 3. All 3 resected cancers (lesions #2, #6 and #7) demonstrated high [^18^F]PARPi uptake at PET and high PARP1 expression on the surgical specimens. The one resected treatment related changes (lesion #5) had low [^18^F]PARPi uptake on PET and also low PARP1 expression in the surgical specimen.

**Figure 3.**
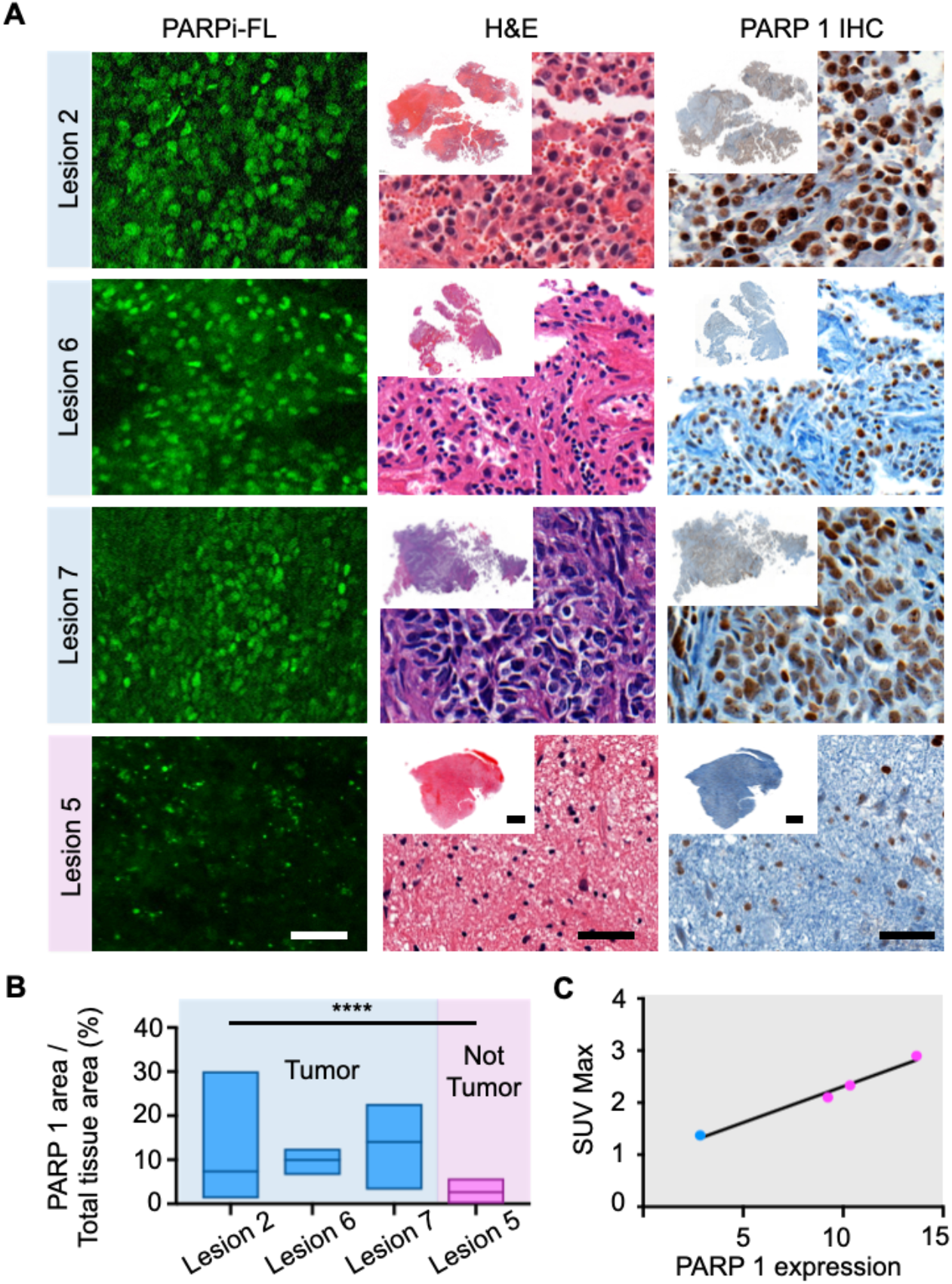
Correlation of PARP1 expression with [^18^F]PARPi uptake. (A) Specific nuclear PARPi-FL uptake was seen in all cancers (lesions 2, 6 and 7). Faint signal was seen in treatment related changes (lesion #5). This differential PARP1 expression in cancer versus treatment related changes was also observed on immunohistochemistry between patients. (B) Differences in quantified PARP1 expression was seen in cancer (lesions #2, 6 and 7 – in blue) when compared to the tissue that only presented treatment related changes (lesion 5 – in pink). (B) Lesion #5, with no viable cancer, had median PARP1 expression over total tissue area of 3%, which was significantly lower than the expression in all cancer specimens, lesions #2, 6 and 7 - 7%, 10% and 14%, respectively, p < 0.001, Kruskal-Wallis test. (C) Correlation of PARP1 expression and the SUV_max_ of [^18^F]PARPi at 60 minutes post injection. Scale bar in slides with high magnification correspond to 50 µm and overviews correspond to 0.5 cm.

Tissue specimens were also stained with the fluorescent version of the PARP inhibitor (PARPi-FL) to confirm uptake. Specific nuclear PARPi-FL uptake was seen in all cancers (lesions #2, #6 and #7). Faint PARPi-FL uptake was seen in treatment related changes (lesion #5) (Fig. 3A), which was verified to be due to PARP1 expression by submitting the tissues to IHC. Lesion #5, with no viable cancer, had a median PARP1 expression over total tissue area of 3% (range, 1 - 5%), which was significantly lower than the expression in all cancer specimens, lesions #2, #6 and #7. The median PARP1 expression over total tissue areas of those lesions were 7% (range 3 - 14%), 10% (range 7 - 10%) and 14% (range 11 - 16%), respectively, p < 0.001, Kruskal-Wallis test (Fig. 3B and Supplementary Table 2). The expression of PARP1 appears to strongly correlate with SUV_max_ uptake values measured with [^18^F]PARPi.

Although the analysis encompasses a small number of data points (only 4 tissue specimens were available), a strong correlation was observed in between PARP1 expression and the SUV_max_ at 60 minutes on the PET scan (Fig. 3C). Notably, with few observations for this analysis, lesion #7 was the influential point driving the value.

#### Intratumoral heterogeneity was demonstrated by different PARP1 expressions

Patient #4 (lesion #6) was scanned with [^18^F]PARPi at 5 months after treatment with laser interstitial thermal therapy (LITT). The surgical specimen from this lesion consisted of 25% cancer and 75% reactive treatment related changes with gliosis and histiocytes (Fig. 4A). The specimen showed greater PARP1 expression in the area of active cancer when compared to the area of treatment related changes, with an average PARP1 expression over total tissue area of 9.33 ± 1.9% versus 0.14 ± 0.06%, respectively. (Fig. 4B). This patient also presented with areas of high and low [^18^F]PARPi uptake on the PET/MR similar to the observed differences in PARP1 expression (Fig. 4C).

**Figure 4.**
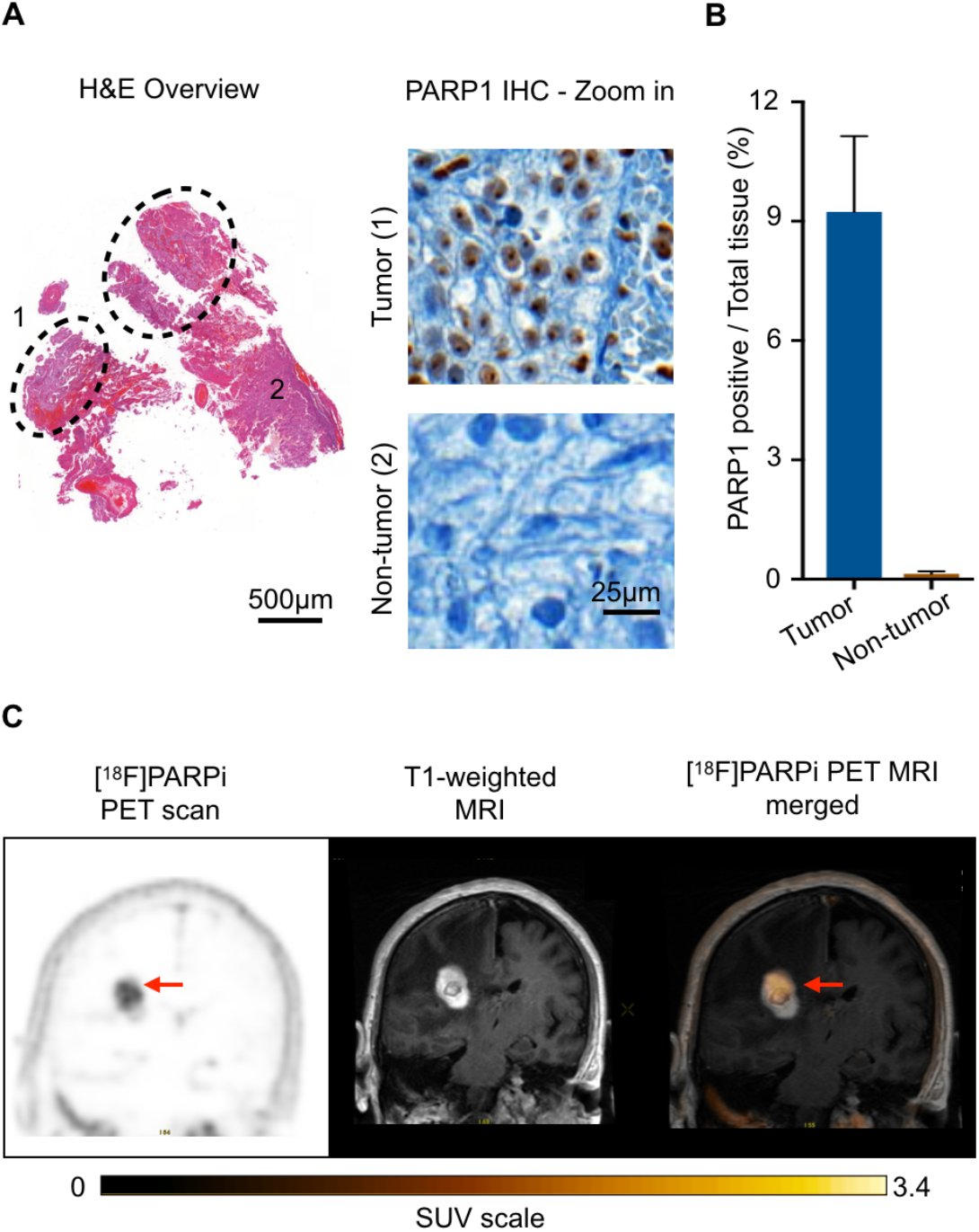
Histology demonstrates [^18^F]PARPi imaging correlation with PARP1 expression. (A) Surgical specimen received from lesion 6 (metastatic renal cell carcinoma status post laser interstitial thermal therapy) was composed of a cancer area represented inside the dotted area with the number 1, and reactive tissue with gliosis and histiocytes represented outside the dotted area by the number 2. PARP1 immunohistochemistry was carried out on the slide and the stain was detected mainly in the cancer areas. (B) Quantification of PARP1 expression over the total tissue area demonstrated that the areas of the specimen composed by cancer had a substantially higher expression of PARP1 when compared to the areas with reactive changes due to treatment. (C) Coronal PET/MR images of the parietal lobe taken with [^18^F]PARPi tracer. Difference in uptake seen on imaging (arrow points to high uptake) is believed to be due to difference in PARP1 expression seen at histology between areas of cancer and areas of treatment change.

#### Fluorescent version of the drug confirms specificity

To demonstrate that [^18^F]PARPi was specific to cancer and not just crossing a permeable blood-brain barrier, we stained (blinded from the final histopathological result) biospecimens received from surgery with the fluorescent version of the compound (PARPi-FL) and imaged under a fluorescence confocal microscope (Fig. 3). PARPi-FL uptake was blocked when tissues were co-incubated with 100-fold of olaparib together with PARPi-FL (Fig. 5A). Quantification of nuclear accumulation was carried out by the presence of PARPi-FL fluorescence signal inside the nucleus of cells. In the unblocked tissue (2,345 nuclei analyzed), 85% of cells showed PARPi-FL uptake whereas in the blocked tissue (1,359 nuclei analyzed), this value was reduced to only 15%. This difference was statistically significant (p < 0.001), confirming blockade of PARPi-FL specific uptake by saturating the PARP1 enzyme with excess of PARP inhibitor (Fig. 5B and Supplementary Table 3).

**Figure 5.**
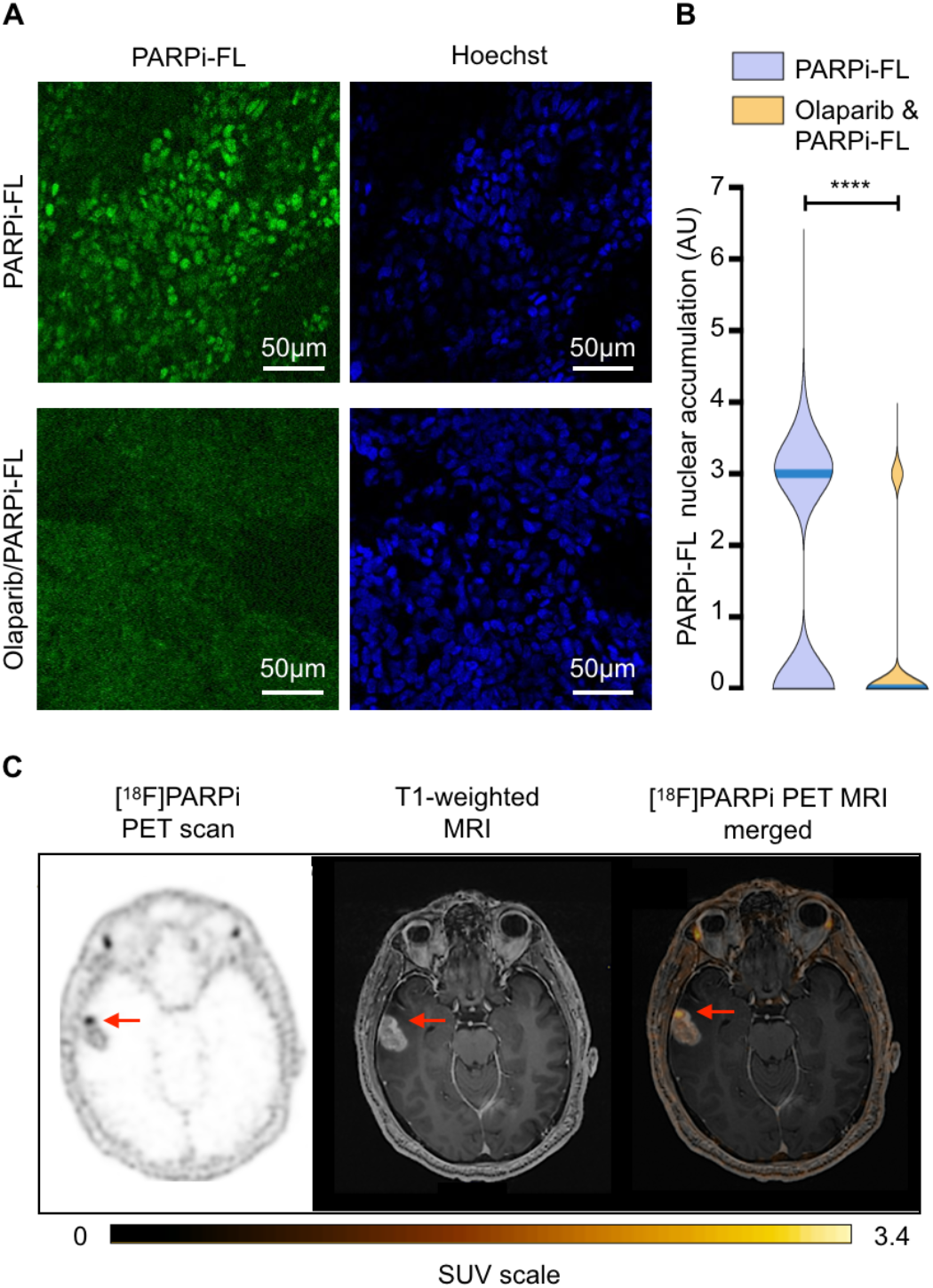
Biospecimen and imaging of lesion #7, untreated glioblastoma. (A) PARPi-FL uptake blocking demonstrates specificity of the compound. Biospecimen stained with the fluorescent version of the compound (PARPi-FL, top row) and blocked (co-incubated with 100-fold excess of olaparib, bottom row). (B) Quantification of nuclear accumulation of PARPi-FL showed a median fluorescence signal significantly higher (p < 0.001) than the mean fluorescence signal edited by the blocked tissue. (C) Axial [^18^F]PARPi uptake map, contrast T1-weighted image, and [^18^F]PARPi map overlaid on contrast T1-weighted image show untreated enhancing cancer in the temporal lobe with high [^18^F]PARPi uptake (arrow).

#### [^18^F]PARPi Metabolism

Research blood draws were obtained from 3 patients, each at 5 timepoints (1-3 minutes, 6-7 minutes, 26-39 minutes, 91-106 minutes and 163 minutes). Using a 2-phase decay curve, we determined the weighted blood half-life to be 1.98 minutes. At 30 minutes, and with decreasing blood pool concentration of the injected tracer, we detected a radiometabolite with a retention time of 13.5 - 16.5 minutes (58 ± 7%) (Supplementary Fig 1A).

#### [^18^F]PARPi has lower normal tissue uptake when compared to [^18^F]FDG

Patient #2 also had a PET/CT using the standard of care radiotracer [^18^F]FDG. Cancer detection with [^18^F]PARPi is not based on metabolic activity, and therefore presented much lower nonspecific uptake in normal brain tissue (Supplementary Fig. 2).

## DISCUSSION

This study describes the cancer-related localization of [^18^F]PARPi in mouse glioma models as well as in first-in-human-brain-cancer patients. Increased [^18^F]PARPi uptake on PET was correlated with increased PARP1 expression of the brain cancer biospecimens. Ancillary [^18^F]PARPi kinetic modeling, PARPi-FL biospecimen imaging and PARP inhibitor blockade confirmed that the uptake was cancer specific.

Modern response criteria such as the Response Evaluation Criteria in Solid Tumors (RECIST), Response Assessment in Neuro-Oncology (RANO) and RANO-brain metastasis (RANO-BM) rely on changes in enhancing size lesions to determine treatment efficacy or failure (*14-16*).

Despite the ubiquity of these and other similar standardized criteria in clinical trials, as well as their growing adoption in clinical practice, there is recognition of the need for advanced imaging techniques to complement these size measurements in treated cancers (*17*). After radiation therapy, chemotherapy and/or immunotherapy, treatment related changes may occur with growing and/or new enhancing lesions that represent inflammatory-mediated changes rather than worsening cancers. Such treatment related changes include both early pseudoprogression occurring in less than 3 months or up to 6-12 months, as well as late radiation necrosis occurring many months or years later, which may occur in up to one-third or more of patients with primary and secondary brain cancers (*18-21*).

We investigated a new radiotracer, [^18^F]PARPi, to bridge this clinical problem. Pre-clinical research has demonstrated uptake in mouse glioma models that is strongly correlated with PARP1 expression and high in cancers and low in normal tissues (*5, 22, 23*). Furthermore, limited uptake was seen in experimentally induced radiation necrosis, despite the presence of blood-brain barrier disruption (*5*). Consistent with previous understanding of the mechanism of PARP upregulation in response to DNA damage (*11, 12, 24, 25*), we confirmed in humans that increased [^18^F]PARPi uptake was correlated with active brain cancer and not with treatment related changes. This has significant implications for patient care, because the accurate and timely non-invasive diagnosis of those changes remain a remarkable imaging challenge. Patients with confirmed recurrent or progressive cancer should stop their current ineffective treatment and instead be considered for possible further resection, radiation therapy, chemotherapy and/or immunotherapy including clinical trials. In contrast, patients with confirmed treatment related changes should continue their current effective treatment and receive supportive care such as additional steroid therapy, and not embark on more aggressive invasive or investigational therapies. The correlation between [^18^F]PARPi PET uptake and PARP1 expression confirms its importance in maintaining genome stability and apoptosis in glioblastoma (*8*), and suggests implications for informing treatment decisions.

[^18^F]PARPi PET also presents the potential for the noninvasive *in vivo* prediction of drug efficacy. Pre-therapy scans already play a role in certain cancers, such as I-123 pretherapy scans in differentiated thyroid cancers before I-131 radioablation (*26*). Given specific localization uptake in brain cancers, [^18^F]PARPi avidity may be useful to quantify PARP1 upregulation in cancer and subsequent sensitivity to systemic PARP inhibitor therapy. This is relevant for brain cancers as PARP inhibitors are being investigated in several ongoing clinical trials (NCT03150862). Besides this, PARP inhibitors are already approved by the U.S. Food and Drug Administration (FDA) for treatment of solid cancers such as ovarian cancer (*27*). PARP inhibitor proof of drug target engagement has been also demonstrated by PET in pre-clinical mouse xenograft models of small cell lung and ovarian cancers (*28, 29*). Demonstrating avid [^18^F]PARPi PET uptake may provide critical data to identify cancers receptive to subsequent PARP inhibitory therapy, versus cancers that may be resistant. Moreover, there are exciting efforts to develop targeted therapeutic options by adding radiotoxic isotopes directly to the olaparib inhibitor scaffold. A recently published study demonstrated promising results using a theragnostic Auger emitting PARP inhibitor (123I-MAPi) in a preclinical model. Taking advantage of the physical properties of Auger emission, dependent on the proximity of the electron emitter to the DNA to cause cellular damage, along with the biological expression of PARP1/2, much higher in cancer when compared to normal cells, it was possible to achieve lethal cancer doses with limited normal tissue toxicity (*30*).

Consistent with the role of PET as an imaging problem solver, the cancer specific avidity of [^18^F]PARPi may also play a role in the differential diagnosis of newly diagnosed brain masses given the apparent lack of correlation with blood-brain barrier disruption that underpins many enhancing brain lesions. The avidity and thereby specificity of the [^18^F]PARPi tracer in cancer-mimicking conditions such as tumefactive demyelinating disease, infarction and abscess, however, is still unknown warranting further research.

One potential limitation of the study is the small human cohort size. Nevertheless, the results of this prospective, first-in-human-brain-cancer study suggest safety as well as a potential role for [^18^F]PARPi PET imaging to discriminate between active brain cancer and treatment related changes, which represents a significant clinical challenge for many patients. A second potential limitation is the absence of histopathology confirmation of cancer versus treatment related changes for 3 of the 7 lesions, in which the outcomes were determined by follow up per standardized response criteria augmented by advanced imaging results. This reflects the realities of clinical care, in which repeat surgery for resection of questionable lesions may not be advisable or feasible in all patients. The 4 resected lesions demonstrated excellent correlation between [^18^F]PARPi uptake and PARP1 expression on IHC, however, as did the advanced imaging results for all lesions.

In conclusion, we present complementary preclinical data and first-in-human-brain-cancer data demonstrating that [^18^F]PARPi uptake is specific to cancer and correlated with PARP1 expression. Although larger trials are necessary, we suggest a role for [^18^F]PARPi in monitoring of patients with the common clinical dilemma of recurrent or progressive brain cancer versus treatment related changes, and future theragnostic applications for both systemic and combination radiotracer-treatment agents.

## MATERIALS AND METHODS

### Preclinical Radiochemistry

[^18^F]PARPi was synthesized using an optimized labeling procedure according to previously described methods (*6, 31, 32*). The preclinical synthesis differs from the clinical synthesis in two ways: (1) [^18^F]fluoride was eluted with a 2 mL solution of K_222_/K_2_CO_3_ (Kryptofix [2.2.2] (4,7,13,16,21,24-hexaoxa-1,10-diazabicyclo [8.8.8]hexacosane (22.5 mg)), 0.02 mL 5 M K_2_CO_3_ and 4% MeCN in H_2_O in V_total_ = 5 mL) and (2) [^18^F]PARPi was isolated by preparatory high-performance liquid chromatography (HPLC) using a flow-rate of 3.0 mL minutes^-1^ and using isocratic 30% acetonitrile in 0.1% triofluoroacetic acid (TFA) solution as the mobile phase. The radiochemical purity showed > 98% (tR = 31 minutes) and the molar activity was 37,000 MBq/µmol (1.0 Ci/µmol).

### Animal Work

Brain cancer development was modeled in p53 deficient nestin/tv-a mice (ntv-a/p53^fl/fl^ mice) using a glioma model based on the RCAS/tv-a system (*33, 34*). All mouse experiments were performed in accordance with protocols approved by the Institutional Animal Care and Use Committee of Memorial Sloan Kettering Cancer Center (MSK) and followed National Institutes of Health (NIH) guidelines for animal welfare. Mice started showing symptoms around 4–5 weeks post-inoculation. To determine the localization of [^18^F]PARPi in the brain, we compared its distribution to FITC-Dextran (Life Technologies, ThermoFisher Scientific, Waltham, MA), which does not cross the blood-brain barrier. We co-injected 150 – 170 µCi of [^18^F]PARPi and 10 kDa FITC-Dextran via the tail vein in cancer-bearing mice.

Animals were sacrificed 1 hour post injection, brains were extracted, frozen and sectioned. Coronal cryosections were exposed to a storage phosphor autoradiography plate (Fujifilm, BAS-MS2325, Fiji Photo Film) overnight at −20°C for radiotracer localization. Adjacent sections were co-stained with Hoechst 33342 and scanned for FITC/Dextran accumulation using a MIRAX scanner. Images were compared using ImageJ to analyze distribution of FITC-Dextran and [^18^F]PARPi in the cancer area (*35*).

### Clinical Radiochemistry

[^18^F]PARPi was produced under good manufacturing practice (GMP) conditions at the MSK Radiopharmacy under investigational new drug (IND) #139,974 similar to previously reported procedures (*25*). Briefly, to obtain [^18^F]PARPi, cyclotron-produced fluorine-18 was trapped on the QMA SEP pack, eluted with 4 mg of tetraethylammonium bicarbonate into a reaction vial containing a teflon magnetic stir bar, and dried by being heated at 90 °C under vacuum. Then, 2 mg of ethyl-4 nitrobenzoate, dissolved in 0.2 mL dimethyl sulfoxide (DMSO), was added to the reaction vessel, and the reaction was heated to 150 °C for 15 minutes and allowed to cool before 50 µL of 1 M NaOH was added, followed by 50 µL of 1 M HCl, yielding 4-[^18^F]fluorobenzoic acid. Thereafter, 4 mg of 4-(4-fluoro-3-(piperazine-1-carbonyl)benzyl) phthalazin-1(2H)-one in 100 µL dry of DMSO were added followed by 10 mg of HBTU dissolved in 100 µL of DMSO and 20 µL of Et_3_N. [^18^F]PARPi was isolated by preparatory high performance liquid chromatography (HPLC), with isocratic 35% acetonitrile in 0.1% TFA solution as the mobile phase, Phenomenex; 6-Phenyl, 5 µm, 250 × 10 mm column as stationary phase at a flow rate of 4.0 mL/minute, and a ultraviolet (UV) detector wavelength of 254 nm. The HPLC system used was a Shimadzu Prominence 20 Series, equipped with a UV detector and a Flow-Ram sodium iodide radioactivity detector supplied by Lablogic, UK. The radiochemical purity range in the [^18^F]PARPi manufactured batches was > 99% (n = 5, t_R_ = 20.2 minutes). The [^18^F]PARPi administered dose ranged from 347.25 to 391.46 MBq (9.385 –10.58 mCi - Specific Activity A_m_ = 105.45 - 461.76 GBq/µmol - 2.85–12.48 Ci/ µmol). It was administered as an I.V. bolus diluted in 6% Ethanol in Saline (0.96 – 2.0 mL). The total injected mass dose (microdose) was 1.06-6.2 µg (2.17 – 12.69 nmol) of [^18^F]PARPi.

### Study Design

This prospective single center, investigator initiated pilot study (ClinicalTrials.gov NCT04173104) examined [^18^F]PARPi PET/MR in patients with brain cancers. The primary objective was to determine [^18^F]PARPi uptake in cancers and treatment related changes. The study was performed according to the Declaration of Helsinki and Good Clinical Practice (GCP) guidelines, was compliant with the Health Insurance Portability and Accountability Act regulations and approved by the local Institutional Review Board and Privacy Board. The authors all vouch for the accuracy and completeness of the data and analyses and for the adherence of the study to the protocol. All patients provided written informed consent before enrollment.

### Patient Selection

The inclusion criteria for the study were patients harboring new or suspected recurrent brain cancer(s) with enhancing lesion(s) ≥ 1.5 cm in diameter. The patients were also required to be ≥ 18 years of age, able to undergo PET/MR scanning, and to receive intravenous gadolinium contrast. All women of childbearing age included in the study had a negative pregnancy test < 2 weeks prior to enrollment. The exclusion criteria were any contraindication to 3T MR scanning per departmental criteria, patient decision to withdraw from the study or failure to comply with protocol requirements. Cohort enrollment was halted at n=5 patients due to the COVID-19 pandemic induced interruption of all non-therapeutic clinical trials in early 2020.

### Patient Imaging

All scans were performed on a 3T PET/MR scanner (Signa, GE Medical Systems, Milwaukee, WI) with lutetium-based scintillator crystal arrays and silicon photomultiplier detectors integrated into the MR gantry to enable simultaneous PET and MR acquisition. PET images were acquired, after peripheral – usually the cephalic vein – administration of 10 mCi of [^18^F]PARPi, with dynamic acquisition for 60 minutes, and additional static 10 minutes acquisitions at 60 minutes and 120 minutes. Volumes-of-interest (VOIs) for SUV measurements were manually placed by an experienced radiologist on lesions visualized by PET images and guided by co-registered MR images, then internally-contoured to select only lesion voxels above a visually guided threshold intensity to select all tracer-avid portions of the lesion for SUV measurements. These SUV measurements were obtained at 60 minutes (SUV_60mean_) and at 120 minutes (SUV_120mean_), as well as ratios normalized to the confluence of the cerebral venous sinuses (ratio SUV_60mean_ and ratio SUV_120mean_). MR sequences were acquired using a 32-channel head coil without and with gadolinium contrast (gadobutrol 0.1 mmol/kg, max 10 mL, Bayer Healthcare, Whippany, NJ) according to the standardized brain cancer protocol (*36, 37*). Per institutional standards, additional advanced imaging also included: 1) Dynamic contrast-enhanced (DCE) T1 perfusion with extended Tofts modeling analysis to calculate K_trans_ and plasma volume maps to determine volume-of-interest ratio measurements for each enhancing lesion (NordicICE 2.3.14, NordicNeuroLab, Bergen, Norway); and 2) Delayed contrast images acquired 60-105 minutes after gadolinium injection with calculation of contrast clearance analysis maps (Elements, Brainlab AG, Munich, Germany) (*38*) and volume-of-interest ratio measurements of voxels demonstrating contrast clearance (encoded in blue, representing cancer) divided by voxels demonstrating contrast clearance and contrast accumulation (encoded in red, representing treatment related change) for each enhancing lesion (software provided by Yael Mardor, PhD, Sheba Medical Medical Center, Tel-Hashomer, Israel).

### Patient Lesion Outcomes

Surgical resection was performed when clinically indicated as per the standard of care. One patient declined cancer resection and clinical and radiological outcomes were used as a surrogate for diagnosis. Resected specimens were classified as cancer when viable cancer tissue was present, and treatment related changes when no viable cancer tissue was present. If histology was not available, due to non-surgical treatment, then lesion outcomes were determined by clinical and imaging follow up based on the Response Assessment in Neuro-Oncology (RANO) criteria for primary brain cancers and the RANO brain metastasis (RANO-BM) criteria for secondary brain cancers (*15, 16, 39*). In patients with brain metastases, maximum 5 enhancing lesions ≥ 1.5 cm in diameter were measured per patient. Per RANO and RANO-BM, respectively, progressive disease (PD) was defined as ≥ 25% increase in the product of perpendicular diameters or ≥ 20% increase in the sum of longest diameters or clinical worsening; partial response (PR) as ≥ 50% decrease in the product of perpendicular diameters or ≥ 30% decrease in the sum of longest diameters; complete response (CR) as the disappearance of all enhancing lesions; and stable disease (SD) as all other conditions. PR and CR required sustained effect for ≥1 month.

### Blood Time Activity Curves

Blood samples for tracer concentration and metabolite analysis were collected. Blood draws were obtained for 3 patients at 5 timepoints after tracer injection, activity counted, and metabolites analyzed (Fig. S1).

### Histopathological Assessment

Lesion outcomes were determined by histopathology when available (n = 4). All cases were reviewed by an experienced neuropathologist. The presence of any viable cancer was considered cancer, and percentages were also recorded when possible. One tissue included both areas of cancer and areas of treatment related changes, which were quantified separately.

### PARPi-FL Synthesis for fresh tissue staining

PARPi-FL was synthesized according to our previously described procedure (*40-42*). Briefly, fluorescent dye BODIPY-FL NHS-ester (1.0 equivalent) was conjugated to 4-(4-fluoro-3-(piperazine-1-carbonyl)benzyl) phthalazin-1(2H)- one (1.0 equivalent) in the presence of Et_3_N (5.0 equivalents) in acetonitrile for 4 h at room temperature. It was purified by preparative HPLC (Atlantis® T3 5 µm column 4.6 × 250 mm, 1 mL/minute, 5 to 95% of acetonitrile in 0.1% TFA in 15 minutes) to afford PARPi-FL in 70–79% yield as a red solid. Analytical HPLC analysis (Waters’ Atlantis T3 C18 5 µm 4.6 × 250 mm column) showed high purity (> 99%, t_R_ = 13.9 minutes) of the imaging agent. The identity of PARPi-FL was confirmed using electrospray ionization mass spectrometry (ESI-MS) (MS(+) m/z = 663.63 [M + Na]^+^) (*13*).

### PARP1 Immunohistochemistry

Paraffin-embedded slides were processed at the molecular cytology core facility using previously described protocols (*17*). Briefly, Anti-PARP1 rabbit monoclonal antibody (46D11, Cell Signaling Technology, Danvers, MA) (0.4 µg/mL) was incubated for 5 h, followed by 1 h of incubation with biotinylated goat anti-rabbit IgG (PK6106, Vector Labs, Burlingame, CA) at a 1:200 dilution. Slides were further scanned (Mirax, 3DHISTECH) to allow for digital histological correlation. Quantification of PARP1 was carried out on digitalized slides. Thresholding was performed (Fiji, Image J) on brown, PARP1 marked with 3’-Diaminobenzidine (DAB) and blue (entire tissue) areas and the relative PARP1-positive area was calculated by dividing the brown (DAB) area by the blue (total tissue area). The entire image slide was encompassed for analysis.

### H&E Staining

Paraffin-embedded slides were processed at the MSK molecular cytology core facility using previously described protocols (*43*). Slides were scanned (Mirax, 3DHISTECH, Budapest, Hungary) to allow for digital histological correlation.

### Tabletop Confocal Microscopy

Freshly excised brain cancers obtained from the patients (n = 4) after surgery (whole-mount) were stained with a solution of 100 nM of PARPi-FL in 30% polyethylene glycol in phosphate buffered saline (PEG/PBS) for 5 minutes, as previously described (*11*). For the blocking experiment, tissues were co-incubated with 100-fold of olaparib together with PARPi-FL. Nuclei were stained with a solution of 10 µg/mL of Hoechst 33342 in PBS. Images were acquired with a laser scanning confocal microscope (LSM880-Live, Zeiss, Germany) using 488 nm laser excitation for PARPi-FL (green), 405 nm for Hoechst (blue) and 561 nm (red) for autofluorescence. Quantification of intensity of PARPi-FL signal was calculated using Fiji (ImageJ) (*12*) by placing the region of interest on the Hoechst nuclear stain and calculating the signal emerged in that same area using the green channel. Nuclear accumulation of PARPi-FL was calculated using arbitrary units (A.U.).

## Statistical Analysis

Statistical analysis of the tissue biospecimens were performed using GraphPad Prism 8 (GraphPad Software, San Diego, CA) and R v3.6.0 (R Core Team (2018), R Foundation for Statistical Computing, Vienna, Austria). Data points represent median values, and error bars represent standard deviations. Statistical analyses for the PARP1 expression on IHC were performed using Kruskal-Wallis test and the correlation with the SUV_max_ on 60 minutes using the Spearman correlation coefficient. Imaging data were examined using Wilcoxon rank sum tests. The blocking tissue experiment results were dichotomized at 3 due to the discrete nature of the data and analyzed with a chi-square test. Statistical significance was determined with p < 0.05. No correction for multiple testing was performed.

## Data Availability

All data associated with this study are present in the paper or the Supplementary Materials.

## REFERENCES AND NOTES

## Acknowledgements

The authors are grateful for the contributions of Matthew Sellitti and the Sloan Advanced Imaging Laboratory (SAIL), and the expert editorial advice from Joanne Chin, Department of Radiology, Memorial Sloan Kettering Cancer Center. We also thank Mark Souweidane (Department of Neurological Surgery, Weill Cornell Medical College) and Melanie Schweitzer (Department of Neurological Surgery, Weill Cornell Medical College) for helpful discussions and assistance with mouse models. The authors gratefully acknowledge the support of the Memorial Sloan Kettering Cancer Center Animal Imaging Core Facility, the MSK Radiochemistry & Molecular Imaging Probes Core, the MSK Molecular Cytology Core, the MSK Nuclear Magnetic Analytical Core and the MSK Center for Molecular Imaging & Nanotechnology.

## Author contributions

RJY and TR conceptualized this study. RJY, PDSF, TR and MPD established methodology and supervised procedures. PLD, GP and SK conducted animal experiments and analysis of the pre-clinical data. PDSF and GP performed PARPi-FL fluorescence and blocking experiments, immunohistochemistry, and fluorescent cell quantification. SR synthetized PARPi-FL and performed blood analysis of the quantified radiolabeled compounds. PDSF, GP, TR and TAB analyzed human tissue specimens. RJY, MPD, AFP and NM organized and collected patient data. RJY, PDSF, PJN, CR, AM, NSM and MPD assisted data including scan investigations. JS developed in house software models. EMB, SR, SKL provided resources. ZZ, AM and MF performed statistical analyses. RJY and PDSF co-wrote the original draft. All authors reviewed and edited the drafts and approved the final manuscript.

## Data and materials availability

All data associated with this study are present in the paper or the Supplementary Materials.

